# Vaccine failure mode determines population-level impact of vaccination campaigns during epidemics

**DOI:** 10.1101/2024.09.30.24314493

**Authors:** Da In Lee, Anjalika Nande, Thayer L Anderson, Michael Z Levy, Alison L Hill

**Affiliations:** Institute for Computational Medicine, Johns Hopkins University, Baltimore, MD; Department of Biomedical Engineering, Johns Hopkins University, Baltimore, MD; Department of Biostatistics, Epidemiology, and Informatics, University of Pennsylvania Perelman School of Medicine, Philadelphia, PA

## Abstract

Vaccines are a crucial tool for controlling infectious diseases, yet rarely offer perfect protection. “Vaccine efficacy” describes a population-level effect measured in clinical trials, but mathematical models used to evaluate the impact of vaccination campaigns require specifying how vaccines fail at the individual level, which is often impossible to measure. Does 90% efficacy imply perfect protection in 90% of people and no protection in 10% (“all-or-nothing”), or that the per-exposure risk is reduced by 90% in all vaccinated individuals (“leaky”), or somewhere in between? Here we systematically investigate the role of vaccine failure mode in controlling ongoing epidemics. We find that the difference in population-level impact between all-or-nothing and leaky vaccines can be substantial when R_0_ is higher, vaccines efficacy is intermediate, and vaccines slow but can’t curtail an outbreak. Comparing COVID-19 pandemic phases, we show times when model predictions would have been most sensitive to assumptions about vaccine failure mode. When determining the optimal risk group to prioritize for limited vaccines, we find that modeling a leaky vaccine as all-or-nothing (or vice versa) can change the recommended target group. Overall, we conclude that models of vaccination campaigns should include uncertainty about vaccine failure mode in their design and interpretation.

## Introduction

Vaccines have been a transformative tool for reducing the burden of infectious diseases around the globe, from early inoculations against smallpox to mass vaccination campaigns for over a dozen pathogens beginning in the 20th century. Since the launch of the World Health Organization (WHO)’s Expanded Immunization Program in 1974, an estimated 154 million lives have been saved and an estimated 9 billion years of life years gained [1] due to vaccines against measles, tetanus, pertussis, tuberculosis, Haemophilus influenzae type B, invasive pneumococcal disease, poliomyelitis, yellow fever, hepatitis B, rotavirus, diphtheria, rubella, Japanese encephalitis, and meningitis A. Another study estimated ∼50 million lives saved by vaccines administered between 2000-2019 [2]. Since the beginning of the COVID-19 pandemic in 2020, dozens of vaccines have been developed and administered to over 5 billion people around the globe [3,4]. In the first year of vaccine administration alone, an estimated 14 million lives were saved due to vaccination [5].

Vaccines stimulate a long-lived pathogen-specific immune response and can act to reduce the probability that vaccinated individuals will acquire infection and transmit it to others, in addition to reducing the likelihood of progressing to severe outcomes if infected. No vaccine is perfect, and the efficacy (if measured in a randomized trial) or effectiveness (if measured in a real-world setting) of a vaccine is defined as the percent reduction in disease occurrence among the vaccinated group compared to the unvaccinated group [6,7]. While vaccine efficacy/effectiveness (“VE”) is an important metric of risk reduction at the *population level*, it does not specify how imperfect vaccines act at the *individual level*. For example, a vaccine that is 80% effective at preventing infection could reduce the per-exposure risk of acquiring infection in each individual identically by 80% (a “leaky” vaccine), or, could provide perfect protection for 80% of the vaccinated population and no protection for the remaining 20% (an “all-or-nothing” vaccine), or, some combination of the two mechanisms [7–9]. It is typically very difficult to measure the distribution of individual protection levels - since doing so requires long-term follow-up with repeated and known exposures (e.g., experimental challenge studies) - and so the mode of vaccine failure is typically unknown. Mathematical models used to predict or assess the success of vaccination campaigns tend to vary in how they encode vaccine-derived protection, but most typically assume only one of these failure modes.

Several mathematical modeling studies have focused specifically on when the mode of vaccine failure can impact predictions about population-level vaccine impact. Early work focusing on the statistical estimation of VE demonstrated that in long-term trials in which a substantial portion of the population is infected during the trial period, the inferred VE can vary based on trial duration and mode of vaccine failure [6,10,11]. This study and more recent work [12] pointed out that imperfect vaccines can be thought of more generally as creating population strata (or a continuum of states) with differential susceptibility. McLean and Blower examined the impact of a hypothetical HIV vaccine administered upon entry into a high-risk community and found that all-or-nothing vaccine-induced protection was generally more impactful, especially over long timescales [13]. Gomes et al. demonstrated that in a simple model of an endemic infection with vaccination administered at birth, the prevalence of infection was lower when vaccine-induced protection was distributed more heterogeneously across the population (i.e., closer to the all-or-nothing extreme) [12]. Similarly, Magpantay et al found that infection levels were lower with all-or-nothing vs leaky vaccines, and that leaky vaccines could lead to a “honeymoon” period after introduction where prevalence is temporarily much lower than the eventual endemic level [14]. Ragonnet et al. examined a model of vaccination at birth for an endemic infection and found that vaccine failure mode, as well as the disease transmissibility and the extent of natural immunity from prior infections, interacted to determine how vaccine efficacy determined population-level impact [15]. Importantly, all of these studies concluded that these differences in population-level impact of vaccination occurred even when the average efficacy, and hence the effective reproduction number under vaccination and the critical vaccination coverage needed for disease eradication, remained the same between vaccine failure modes.

In the case of emerging epidemics like COVID-19, many of the assumptions employed in previous models assessing the impact of vaccine failure mode are less appropriate. For emerging epidemics, vaccines are typically administered to large portions of the population at once (not just infants or young children) and in the middle of the outbreak (not before), and the impact of interest is the cumulative portion infected within some time period during the epidemic phase (not the eventual endemic prevalence), and the effect of demographic change (e.g., births and deaths) is often negligible on the timescale of pathogen spread. Thus, there is a need to understand when and how vaccine failure mode influences the population-level effects of vaccination during an epidemic. In this study, we extend previous work to examine how the distribution of individual-level vaccine efficacy influences final epidemic size as a function of timing and speed of vaccine roll-out, overall vaccine coverage, baseline transmission rates, and average vaccine efficacy.

## Methods

### Modeling imperfect vaccines

To systematically examine how vaccine failure mode influences predictions of population-level impacts of vaccination, we used a simple compartmental infectious disease model (Figure 1). In the absence of vaccination, individuals can be classified as susceptible to infection (S), exposed but not yet infectious (E), infectious (I), or recovered and immune (R). We assume that the population is well-mixed, that the total population size is constant, and that demographic events such as births and deaths can be ignored on the timescale of the epidemic. Recovered individuals are assumed to be completely immune to reinfection, and all unvaccinated individuals have equal susceptibility. Vaccination moves susceptible individuals into one of two vaccinated groups: in vaccinated-and-protected individuals (V_R_) the per exposure contact rate is reduced with efficacy ε_L_ (i.e., efficacy of “leaky” protection), while in vaccinated-but-susceptible individuals (V_S_) the per exposure contact rate is unchanged (efficacy of zero). A fraction ε_A_ of individuals receiving the vaccine move to V_R_ while the remaining fraction 1-ε_A_ move to V_S_ (ε_A_ is the efficacy of “all-or-nothing” protection). This model allows us to consider a continuum of scenarios between a leaky vaccine and an all-or-nothing vaccine, while still considering only two strata of vaccinated individuals. The “leaky” extreme of vaccination is represented as 100% of vaccinated individuals moving to the vaccinated-and-protected state (ε_A_ = 1, 0<ε_L_<1), while the all-or-nothing extreme is encoded as 100% protection from infection in the vaccinated-and-protected class (0<ε_A_<1, ε_L_ = 1). A vaccine that acts through some combination of these mechanisms can be represented by intermediate parameter values. Note that throughout the paper, vaccine efficacy is used only to describe the reduction in the per contact risk of infection (e.g., VE against infection), and we ignore additional effects a real-world vaccine might have, such as reducing the risk of symptomatic or severe disease if infected, reducing the duration of infectiousness, reducing pathogen shedding during infection, or synergizing with infection-induced immunity. Thus, we assume that the duration of the latent period (σ) and infectious period (ˠ), the infectiousness of infected individuals (β), and the perfect life-long immunity conferred by infection are all identical regardless of vaccination status at the time of infection.

**Figure 1:**
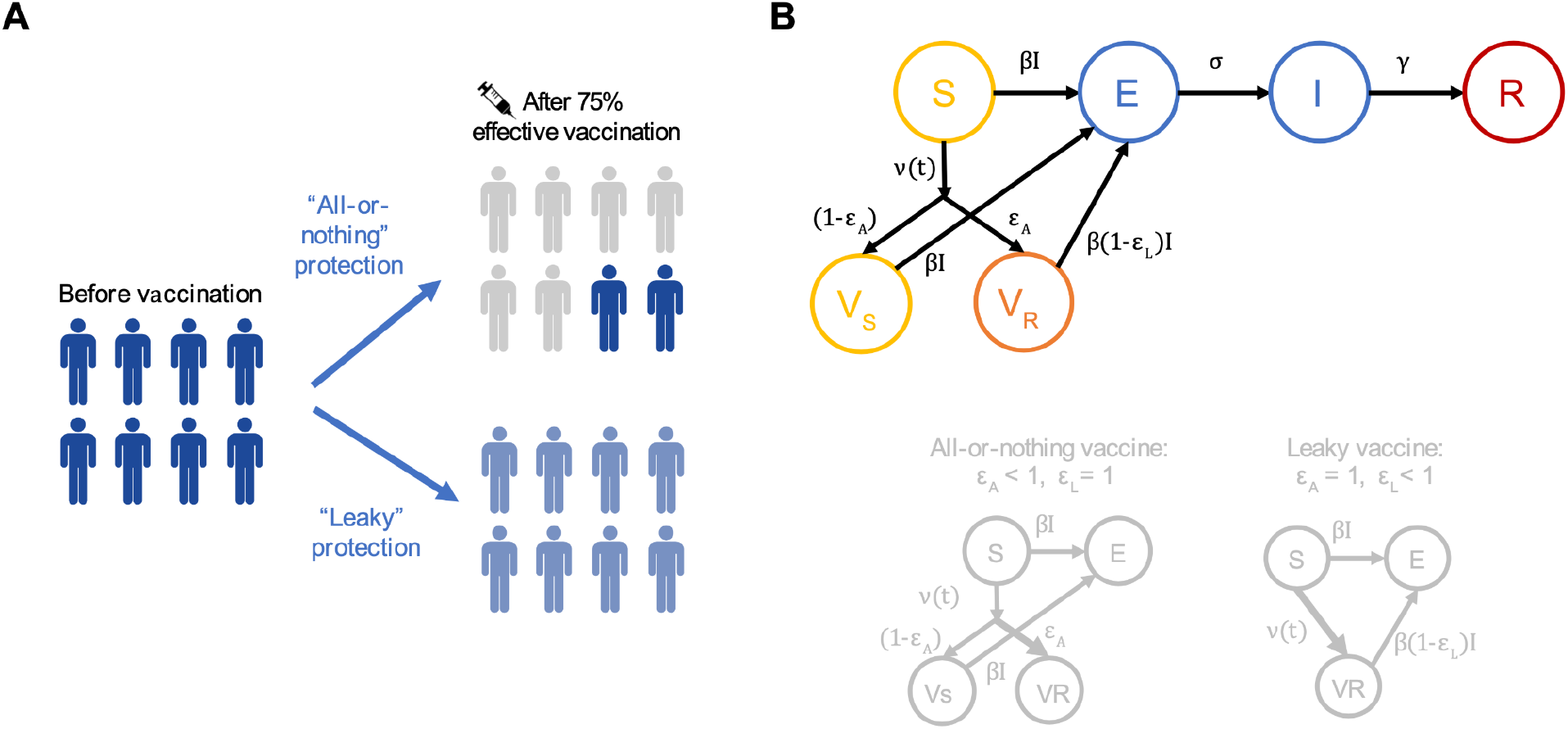
Mechanisms of vaccine failure. A) The two mechanisms of vaccine failure/protection. Shade of blue represents the degree of susceptibility before and after vaccination. With all-or-nothing protection, some individuals are completely protected (gray shade, no remaining susceptibility), while others maintain baseline susceptibility (dark blue). For leaky protection, all individuals experience the same reduction in susceptibility after vaccination (light blue). B) Schematic of the full compartmental model consisting of individuals susceptible to infection (S), infected but not yet infectious (E), infectious (I), recovered and immune (R), vaccinated-and-protected individuals (V_R_), and vaccinated-but-susceptible individuals (V_S_). β is the per-contact-infectiousness of an infected individual, σ is the rate at which infected individuals become infectious (1/σ average duration of latent period), and ˠ is the rate of recovery (1/ ˠ average duration of infectiousness). Vaccination occurs at rate *v*(*t*). ε_A_ is the proportion of individuals protected at all by vaccination, while ε_L_ is the degree of protection in protected individuals. For all-or-nothing protection, ε_L_ = 1 and efficacy is described by ε_A_ with 0<ε_A_<1. For leaky protection, ε_A_ = 1 and efficacy is described by ε_L_ with 0<ε_L_<1.

This model is represented by the following system of ordinary differential equations:

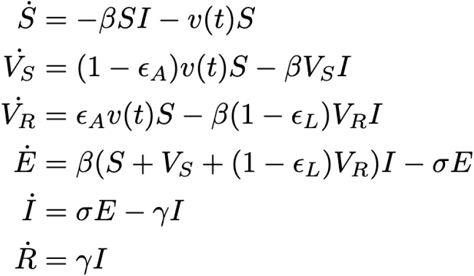

where the state variables S, V_S_, V_R_, E, I, and R are proportions of the population. A vaccination campaign is described by a time dependent vaccination rate *v*(*t*), which describes the proportion of the population vaccinated per unit time. We consider two models of vaccination campaigns (i) a “rapid” vaccination campaign taking place instantaneously at time t_V_ and targeting a fraction f_V_ of the population,

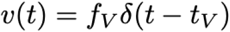

where δ is the Dirac delta function. And (ii) a “rolling” vaccine campaign beginning at time t_V_ at initial rate v_max_ and continuing until a portion f_V_ of the population are vaccinated,

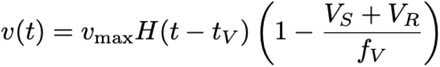

where *H* is the Heaviside step function.

The basic reproduction number for this model, assuming vaccination takes place before the epidemic begins and reaches a total fraction f_V_ vaccinated, is:

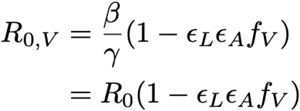

where R_0_ is the value in the absence of vaccination. This results in a critical vaccinated fraction to prevent disease emergence:

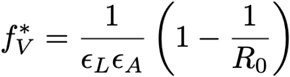

If vaccination begins after the epidemic has begun, then assuming an instantaneous vaccination campaign targeting only susceptible individuals and beginning after a fraction f_R_ have already been infected and are immune to reinfection, the critical vaccination coverage is reduced to

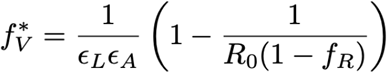

For our study of the effect of different kinds of vaccination, we chose model parameters to correspond roughly with the timescale of COVID-19 transmission.

**Table 1.**
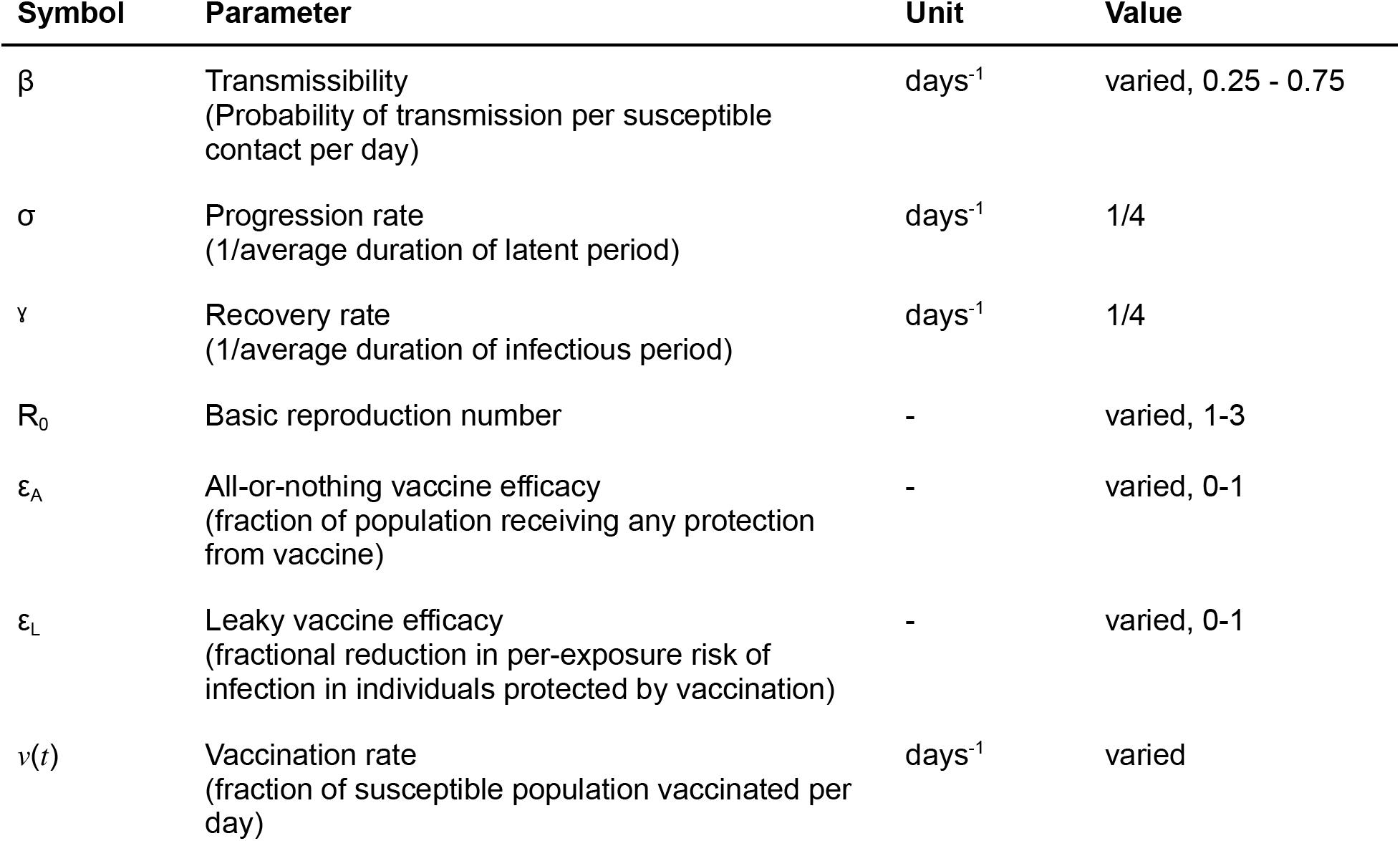
Summary of parameters and symbols used in the model. Values that weren’t varied are those based roughly on COVID-19.

| Symbol | Parameter | Unit | Value |
| --- | --- | --- | --- |
| $\beta$ | Transmissibility<br>(Probability of transmission per susceptible contact per day) | days<sup>-1</sup> | varied, 0.25 - 0.75 |
| $\sigma$ | Progression rate<br>(1/average duration of latent period) | days<sup>-1</sup> | 1/4 |
| $\gamma$ | Recovery rate<br>(1/average duration of infectious period) | days<sup>-1</sup> | 1/4 |
| $R_0$ | Basic reproduction number | - | varied, 1-3 |
| $\epsilon_A$ | All-or-nothing vaccine efficacy<br>(fraction of population receiving any protection from vaccine) | - | varied, 0-1 |
| $\epsilon_L$ | Leaky vaccine efficacy<br>(fractional reduction in per-exposure risk of infection in individuals protected by vaccination) | - | varied, 0-1 |
| $v(t)$ | Vaccination rate<br>(fraction of susceptible population vaccinated per day) | days<sup>-1</sup> | varied |

### Comparing ‘All-or-Nothing’ and ‘Leaky’ Vaccines

In order to systematically compare the two modes of vaccine failure, we focused on nine different general regimes. First, we varied the time at which the vaccine was administered. Vaccination began either (i) at the beginning of the epidemic, (ii) when 10% of the population was immune (R state), or (iii) 25% of the population was immune. Next, for the rapid vaccination model we varied the percent of the population vaccinated and focused on 50%, 75% and 100% coverage levels. For the “rolling”, continuous vaccination model, we fixed the coverage to 75% and instead varied the rate at which susceptible people get vaccinated. We considered 1%, 3% and 5% of susceptible and unvaccinated individuals to get vaccinated daily until the desired coverage is reached.

For each of these nine scenarios, we varied the R_0_ values from 1.0 to 3.0 and vaccine efficacy from 50% to 100%, and the herd immunity threshold 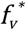 was calculated at each point based on its R_0_ value and vaccine efficacy. The impact of each of the vaccine failure mechanisms (*VI*_*ADN*,_ *VI*_*Leaky*_) were evaluated by calculating the percent reduction of the final epidemic size compared to that of without vaccination. Note, the differences in vaccine impact (Δ*VI*) were calculated by subtracting the percentage reduction of the leaky vaccine from the all-or-nothing vaccine, as the all-or-nothing vaccine demonstrated a greater percentage reduction for all scenarios.

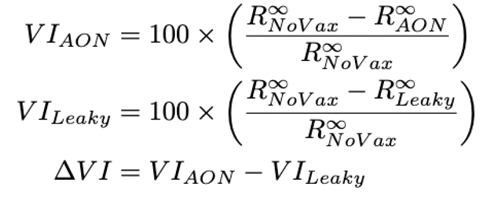

where 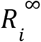 corresponds to the number of cumulative infections at the end of the epidemic (final epidemic size) for the no vaccination, vaccination by all-or-nothing or by leaky vaccines.

Parameter combinations for which the R_0_ value was too low for the disease to reach the target % population immunity before the vaccine campaign started was excluded from the final comparison.

Analysis was done in Python 3.0, and our code is open-source and available on Github: https://github.com/diane-lee-01/VaccineEfficacy

### Model for exploring vaccine allocation decisions

In order to explore how the choice of the vaccine failure mode affects vaccine allocation decisions, we consider a modification of the compartmental model described above – the population is split into two equally-sized groups where the groups can differ in terms of their relative infectiousness, susceptibility, chance of developing severe disease if infected, and vaccine efficacy (see SI Methods for full model equations). All other parameters are the same for each group and there is no other structure in the population (that is, it is fully-connected: individuals are equally likely to have potentially-infectious contacts with other individuals in either population). We assume only one group can be vaccinated (an instantaneous vaccine administered at the start of the epidemic) and for each mode of vaccine failure (‘leaky’ and ‘all-or-nothing’), we calculate the vaccine impact (defined as the percentage reduction of the infected population compared to without vaccination) of vaccinating either of the two groups over a range of parameter values. The group associated with the higher impact of vaccination is considered to be the optimal vaccination choice. We then compare whether this choice varies or stays the same for the two modes of vaccine failure.

We focus on three scenarios where the differences between the groups are associated with trade-offs such that it is unclear which group should be vaccinated to achieve the maximum reduction in the final epidemic size. In all three scenarios, one group is more susceptible, but additionally is either less infectious, experiences lower vaccine efficacy, or has a lower propensity for severe disease if infected. For each of these scenarios we consider three sub-scenarios with different combinations of baseline vaccine efficacies (VE) and coverage-levels: (i) 50% VE, 80% coverage, (ii) 80% VE, 50% coverage, and (iii) 80% VE, 80% coverage. We then explore how the difference in vaccine impact between vaccinating one group versus the other changes in these sub-scenarios by varying R_0_ and the parameters related to the relative differences between the two groups; for example, for the first we vary the relative reduction in infectiousness in one group compared to the relative reduction in susceptibility of the other (see Supplemental Methods for more details). We quantify vaccine impact as percentage reduction of the final epidemic size (in the total population - both groups combined) compared to that of without vaccination. We compare vaccine impact when only one group vs only the other is vaccinated to decide the “optimal” group to vaccinate. However, we assume that if the magnitude of difference in vaccine impact is less than 1%, then it doesn’t matter which group is vaccinated. Finally, we compare these allocation decisions across the parameter space for the two models of vaccine failure, and identify regions where they differ, that is, where the leaky model chooses one group to vaccinate whereas all-or-nothing chooses the other group. Note, for the scenario where groups differ by propensity for severe infection, we focus on the total number of severe infections as the outcome of interest instead of the final epidemic size.

## Results

### When does vaccine failure mode matter?

To understand when the choice of the vaccine failure mode affects model outcomes, we compared predictions for the final epidemic size when we assumed either all-or-nothing or leaky vaccine-induced protection. We found many instances when the two assumptions led to large differences in model predictions. For example, we considered an epidemic spreading with R_0_=2.5 and infecting 90% of the population in the absence of vaccination, and simulated vaccinating 50% of the population prior to the start of the epidemic with either a leaky or all-or-nothing vaccine with 60% efficacy (Figure 2A/B). In the very early stages of the epidemic, we found that vaccination slows down the spread of the epidemic similarly for both types of vaccine failure. However, their effect on the epidemic diverges as population immunity increases. The all-or-nothing vaccine leads to a lower epidemic peak that is reached earlier as compared to the leaky one, and has a lower final attack rate – 50% (all-or-nothing) versus 63% (leaky) of the population infected by the end of the epidemic.

**Figure 2:**
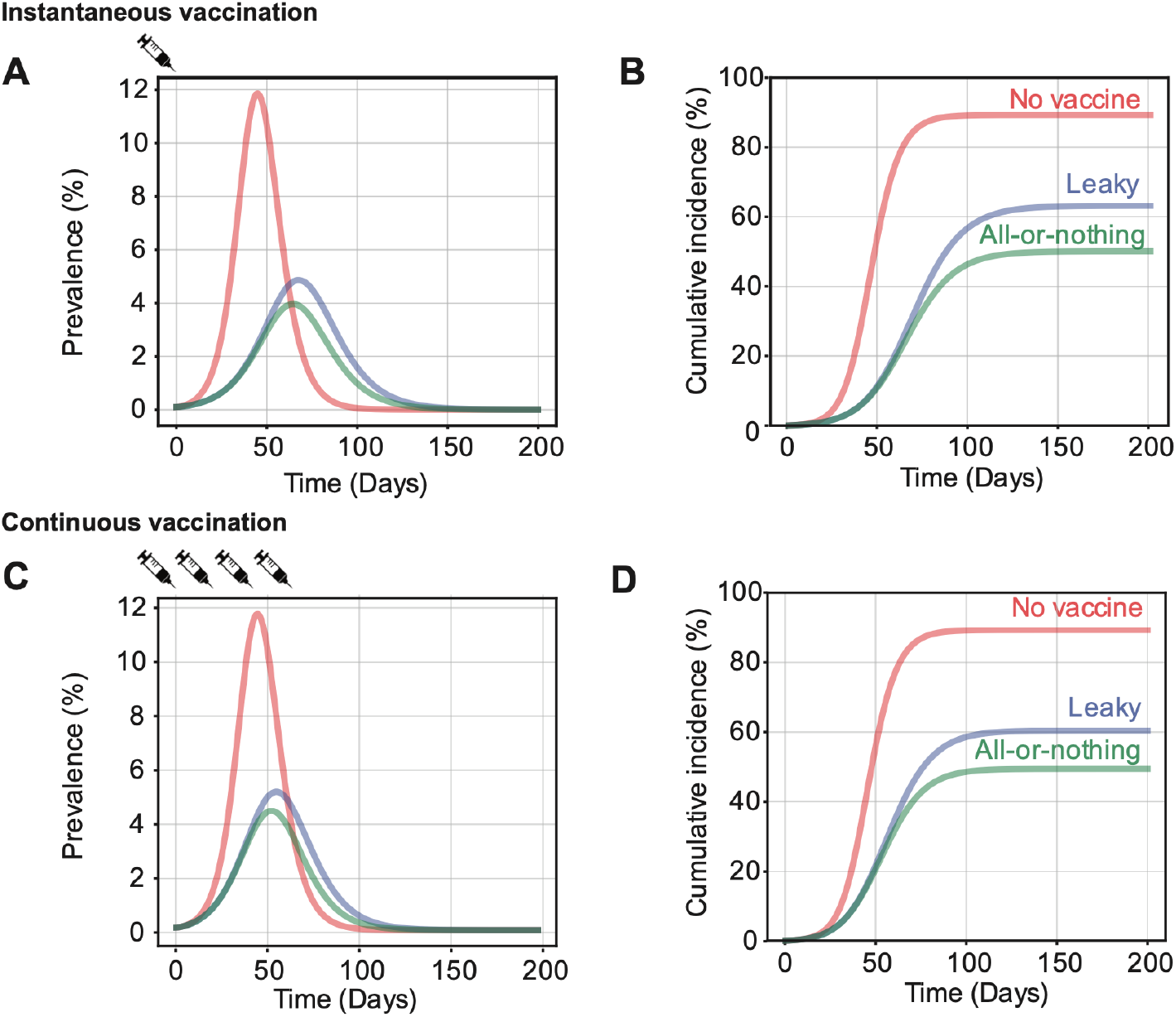
Vaccine failure mode determines impact of vaccination campaign. Time course of infection in the absence of vaccination (red lines), compared to a scenario with administration of a vaccine with either leaky (blue line) or all-or-nothing (green line) mode of protection. For the rapid vaccination scenario (A, B), 50% of the population is vaccinated prior to the start of the epidemic. For the rolling vaccination scenario (C, D), 5% of the susceptible individuals are vaccinated daily after the start of the epidemic until 50% of the population is vaccinated. Results are shown for R_0_ = 2.5, and vaccine efficacy = 60%.

The difference in vaccine impact between the two modes of vaccine failure is highly dependent upon model parameters, such as the timing of vaccine administration, vaccine coverage, vaccine efficacy, and R_0_ in the absence of vaccination (Figure 3). We defined the *vaccine impact* as the reduction in the final epidemic size compared to the scenario without any vaccination, and then we compared the vaccine impact between leaky and all-or-nothing vaccines (see Methods for more details). We find that the impact of all-or-nothing vaccines is always greater than or equal to that of leaky vaccines, with the difference in impact being more significant when the vaccine efficacy is intermediate (∼50%), the epidemic is spreading rapidly (high R_0_), the vaccine is administered before substantial spread of the epidemic, and more people are vaccinated. Overall, the difference in vaccine impact is substantial (≥5%) for a large portion (∼ 35%) of the parameter space we considered, with the maximum difference being around 30% in the final epidemic size.

**Figure 3:**
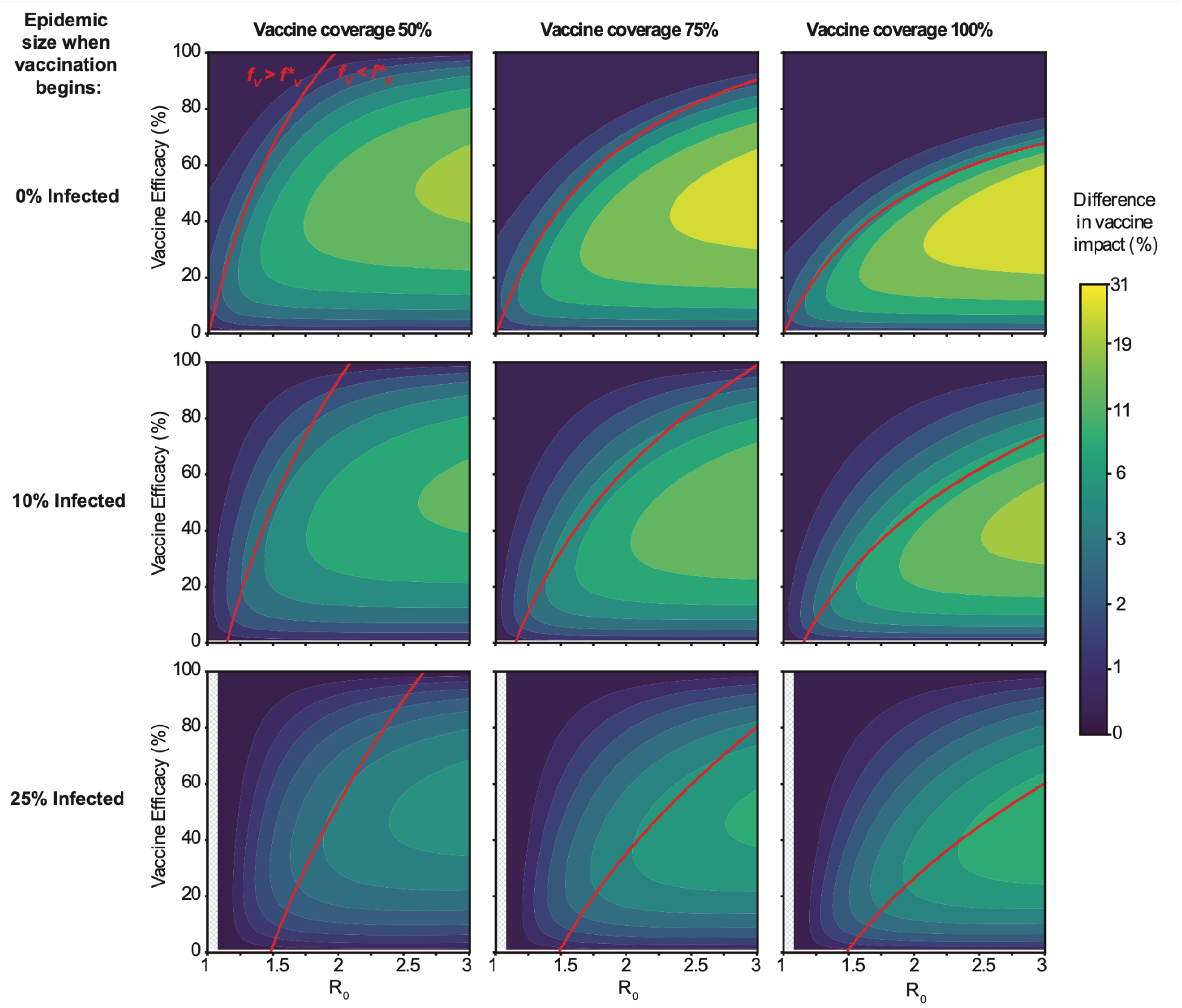
Difference in impact between all-or-nothing and leaky vaccines for rapid vaccine administration. Vaccine impact is defined as the percent reduction in the final epidemic size, compared to a scenario with no vaccination. The heatmap color indicates the difference in vaccine impact between all-or-nothing vs leaky vaccines, for a range of values for disease transmissibility (R_0_, x axis), vaccine efficacy (VE, y axis), timing of vaccine administration (rows, vaccine is administered all at once, at a fixed point in the epidemic), and vaccine coverage (columns, proportion of population vaccinated). The red line separates regions in the parameter space where the fraction vaccinated (*f*_*v*_) is greater (above) or less (below) than the critical vaccination threshold (*f\**_*v*_, aka herd immunity threshold). Uncolored hashed regions correspond to parameter combinations where disease R_0_ was too small for the epidemic to reach the target size for vaccine administration (so vaccine was never administered).

In the previous examples, we approximated a rapid vaccination campaign by assuming all vaccines were administered instantaneously at a fixed time during the epidemic. We also considered more gradual, continuous vaccination campaigns (“rolling vaccination”, Figures 2C,D) and similarly found that vaccine failure mode could influence model predictions. For example, we simulated vaccinating 5% of unvaccinated individuals daily after the start of the epidemic with either a leaky or all-or-nothing vaccine with 60% efficacy until a coverage of 50% was reached. The difference between the two is more pronounced later in the epidemic and the all-or-nothing vaccine had a lower final epidemic size (49%) compared to the leaky vaccine (60%). Considering a wide range of parameter values, we find that the difference in vaccine impact is more sensitive to the rate at which vaccination occurs rather than the final coverage levels (Figure 4, see Suppl. Fig S1). The difference between all-or-nothing versus leaky vaccines is more significant when the vaccine efficacy is intermediate (∼50%), the epidemic is spreading rapidly (high R_0_), the daily rate of vaccination is high, and when the campaign starts earlier in an epidemic.

**Figure 4:**
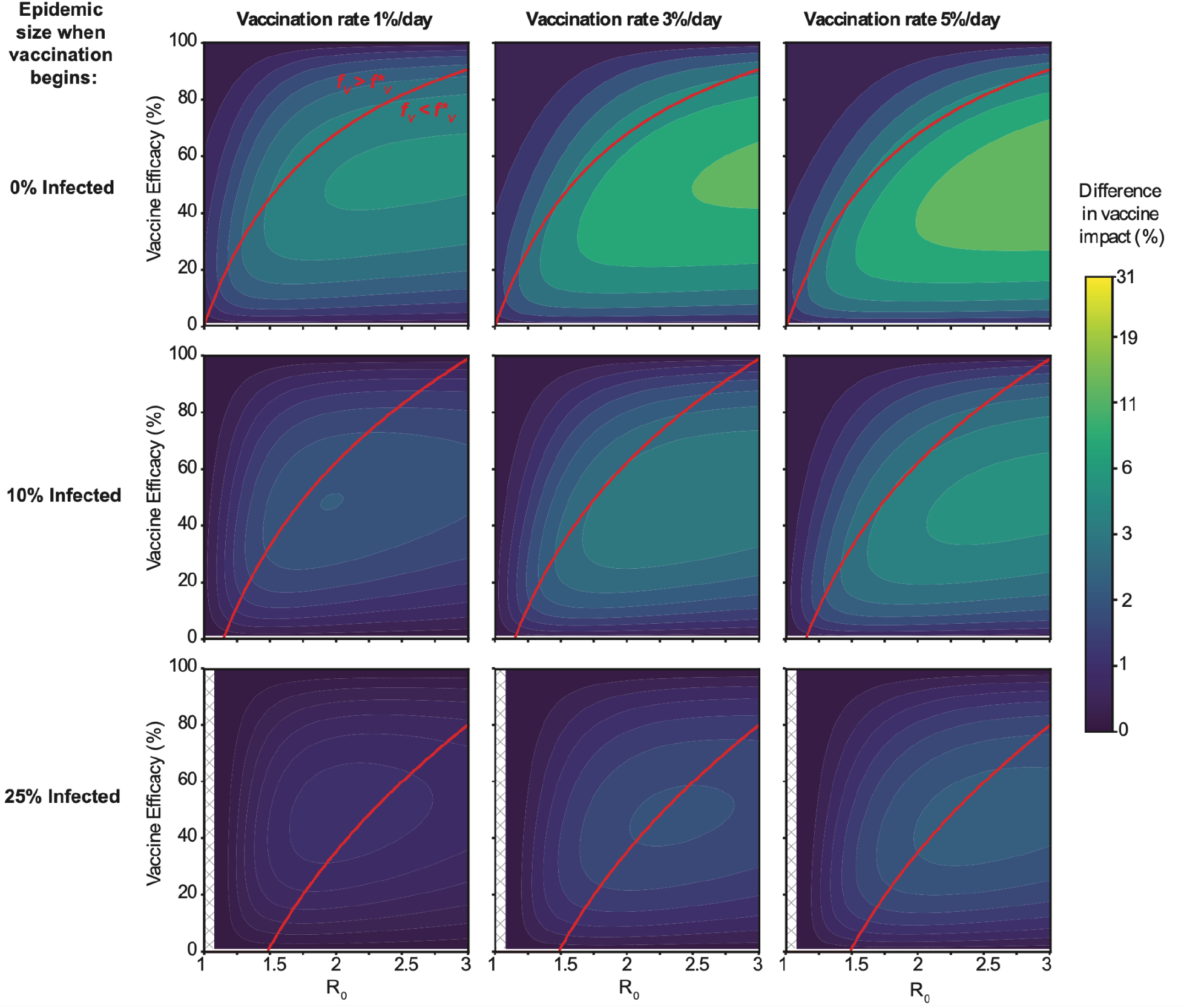
Difference in impact between all-or-nothing and leaky vaccines for rolling vaccine administration. Vaccine impact is defined as the percent reduction in the final epidemic size, compared to a scenario with no vaccination. The heatmap color indicates the difference in vaccine impact between all-or-nothing vs leaky vaccines, for a range of values for disease transmissibility (R_0_, x axis), vaccine efficacy (VE, y axis), timing of beginning of vaccine administration in terms of epidemic size (rows), and speed of the vaccination campaign (proportion of unvaccinated population vaccinated per day). In all cases final vaccine coverage is 75%. See Methods for more details on how rolling vaccination was implemented. Uncolored hashed regions correspond to parameter combinations where disease R_0_ was too small for the epidemic to reach the target size for vaccine administration (so vaccine was never administered).

The difference in population-level protection offered by leaky vs all-or-nothing vaccines with identical efficacy can be explained by comparing breakthrough infections – the number of vaccinated individuals that eventually become infected – between the two models. Both all-or-nothing and leaky vaccines lead to equal instantaneous reduction in population susceptibility by reducing the per-exposure infection risk. However, if the epidemic is not immediately curtailed and continues to circulate in unvaccinated individuals, vaccinated individuals may be exposed multiple times. In the case of all-or-nothing protection, regardless of the number of exposures, only (1-VE)% of vaccinated individuals can ever become infected. In contrast, for leaky vaccines, after multiple exposures it is possible that all vaccinated individuals eventually get infected. Even if vaccine efficacy and coverage is the same in both models, the leaky model is always associated with an equal or a larger number of breakthrough infections as compared to the all-or-nothing model (See SI Methods for derivation). Intuitively, this occurs because for a fixed R_0_ in the all-or-nothing model the average susceptibility of the next infected individual is always greater than in the leaky model and hence susceptibility drains out of the population more quickly leading to a smaller epidemic size. This difference is highest in the regime where multiple exposures are possible (high R_0_) and when vaccine efficacy is intermediate (VE∼50%), because if vaccine efficacy is high, both vaccine types are associated with few breakthrough infections and if vaccine efficacy is low, both are associated with a large number of breakthrough infections. This effect is related to two previous findings in the modeling literature: 1) In clinical trials measuring vaccine efficacy or observational studies of vaccine effectiveness, multiple exposures during the study period could bias estimates of VE downwards for leaky vaccines [8,10,11,16,17], and 2) Populations with high heterogeneity in disease susceptibility (due to natural variation as well as vaccine-induced protection) tend to support smaller epidemics than those with uniform susceptibility [12,18–24].

### Applications to COVID-19 vaccine impact predictions

Throughout the COVID-19 pandemic, models have been used to assess the impact of vaccination campaigns and to guide decisions about optimal vaccine allocation. Very few considered how the choice of modeling vaccine failure might affect conclusions (for some exceptions, see [25–28]). Our results demonstrate that the difference in model predictions between these two assumptions can vary substantially depending upon the nature of the outbreak and the details of the vaccination campaign. Next, we examine some scenarios inspired by real-world vaccination campaigns that occurred during specific waves of the COVID-19 pandemic to assess whether the choice of the mode of vaccine failure would have appreciably affected model outcomes (see Supplemental Methods for details).

First we consider the scenario in which an epidemic wave is rapidly controlled via vaccination, as was seen in Israel in the late fall of 2020 through winter of 2021 when a large wave of SARS-CoV-2 infections emerged just as vaccines became available. We estimate 10% prior immunity to approximate the size of the epidemic prior to the start of their respective epidemic waves (see Supplemental Methods). To roughly recreate the vaccine campaigns in Israel during this time, vaccine administration was assumed to be rolling from the start of the outbreak, and occurred at a rate that ensured ∼20% of the population was vaccinated each month until a total coverage of 60% was reached. R_0_ was chosen such that the effective R_0_ at the start of the outbreak was ∼1.5. Since these waves occurred when the non-variant-of-concern strains of SARS-CoV-2 were circulating (i.e., Alpha), we assume that the vaccine efficacy against any infection was around 90% [29]. In such situations of rapid epidemic control via vaccination, we find that the choice of the vaccine failure model has no observable effect on the predicted final epidemic size, with 37% of the population being infected under both the models (Figure 5, first column).

**Figure 5:**
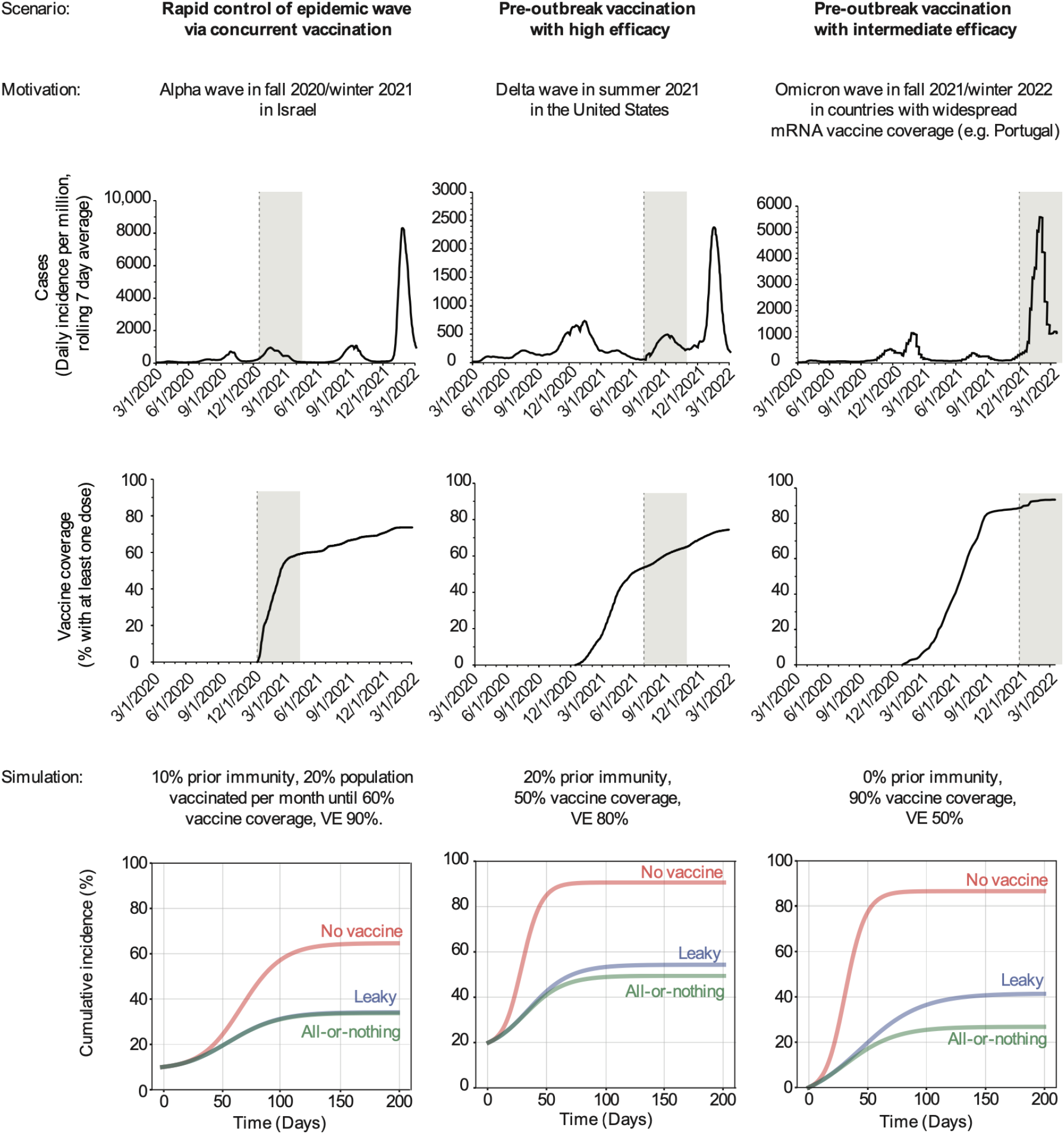
Differences between the two vaccine failure models for scenarios motivated by the COVID-19 pandemic. Each column describes a different setting during the COVID-19 pandemic that motivated the choice of parameters and vaccination campaign for the simulation. Top row: Data, reported COVID-19 cases over time for the country motivating the scenario, with the shaded region in time highlighting the time period of interest. Middle row: Data, reported proportion of the population vaccinated against COVID-19 over time. Bottom row: Simulation results, showing cumulative infections over time in the absence of vaccination (red line) and in the presence of vaccination, where vaccine offers leaky (blue) or all-or-nothing (green) protection. More details on each scenario are provided in the Supplemental Methods.

Motivated by the SARS-CoV-2 Delta variant wave that occurred during the summer of 2021 in the US, our next scenario considers the situation where a highly effective vaccine was administered prior to an outbreak, but without adequate coverage. Here we estimate prior immunity nationally to be 20%, based on seroprevalence of antibodies against SARS-CoV-2 N protein at the beginning of June 2021 [30,31], roughly when the epidemic wave began [32]. We assumed a vaccine coverage of 50% before the wave began, estimated from data on the percent of fully vaccinated individuals in the US by June-July 2021 [3,4], and assumed a vaccine efficacy of 80%, based on mRNA vaccine efficacy against any infection for the Delta variant [29]. We assumed vaccine impact projections were being made early in the wave, when R_0_ was estimated at 3 (1.5 in the presence of infection and vaccine-induced immunity), and assumed no further interventions or pathogen evolution. We find that the choice of vaccine failure mode for such a situation has a small but significant effect on the final epidemic size — 54% of the population is predicted to get infected under the leaky vaccine as opposed to 49% under the all-or-nothing one (Figure 5, middle column). Note that these simulations are not meant to recreate the exact observed epidemic course - but to reflect simplified versions of vaccine impact projections that may have been conducted early on during these waves/vaccine campaigns.

Finally we focus on a situation in which a partially effective vaccine is administered with high coverage before an outbreak. We were motivated by the Omicron variant wave that occurred in the winter of 2021/2022 in countries that had especially high mRNA vaccination coverage; for example, Portugal [4]. Given the immune evasive properties of Omicron, we assumed there to be no prior immunity in the population against infection. We assume vaccine coverage to be 90% which was estimated from data on the percent of fully vaccinated individuals by December 2021 [3], and vaccine efficacy was assumed to be 50% (in reality, studies give a wide range of estimates, and find strong dependence on time since last dose and whether a booster dose was received [33–37]). We chose an R_0_ of 2.3 such that on average about 10% of the population is infected each month until the epidemic ends, roughly based on the initial speed of the Omicron wave. For this scenario, we find there would be large differences in the predicted final epidemic size under the two models of vaccine failure — 41% of the population gets infected under the leaky vaccine as opposed to 27% under the all-or-nothing one (Figure 5, last row).

### Vaccine allocation decisions

Real-world vaccination campaigns face constraints due to limited vaccine supplies, high vaccine costs, and the speed with which they can be administered. Mathematical models are a useful tool to compare different vaccine distribution strategies – such as prioritizing groups based on age, comorbidities, or other risk factors – and to find the one that leads to the largest reduction in population-level disease burden [25,38–41]. To ensure a robust public health response, modelers should characterize the uncertainty in the optimal predicted strategy due to model assumptions. Here we focus on understanding whether and how the assumed mode of vaccine failure affects vaccine allocation decisions generated from models. We create a simple model of a heterogeneous population for which decisions about vaccine allocation may be debated. We consider a well-mixed population divided into two groups of equal size that may differ in their infectiousness, susceptibility, vaccine efficacy and disease severity (Figure 6, see Methods for more details). We assume only one group can be vaccinated, and use our model to determine which allocation strategy (Group 1 or Group 2) would result in the largest reduction in the final epidemic size. We consider three scenarios where the differences in disease risk between the groups is such that without the aid of a model, there is ambiguity about which group should be vaccinated.

**Figure 6:**
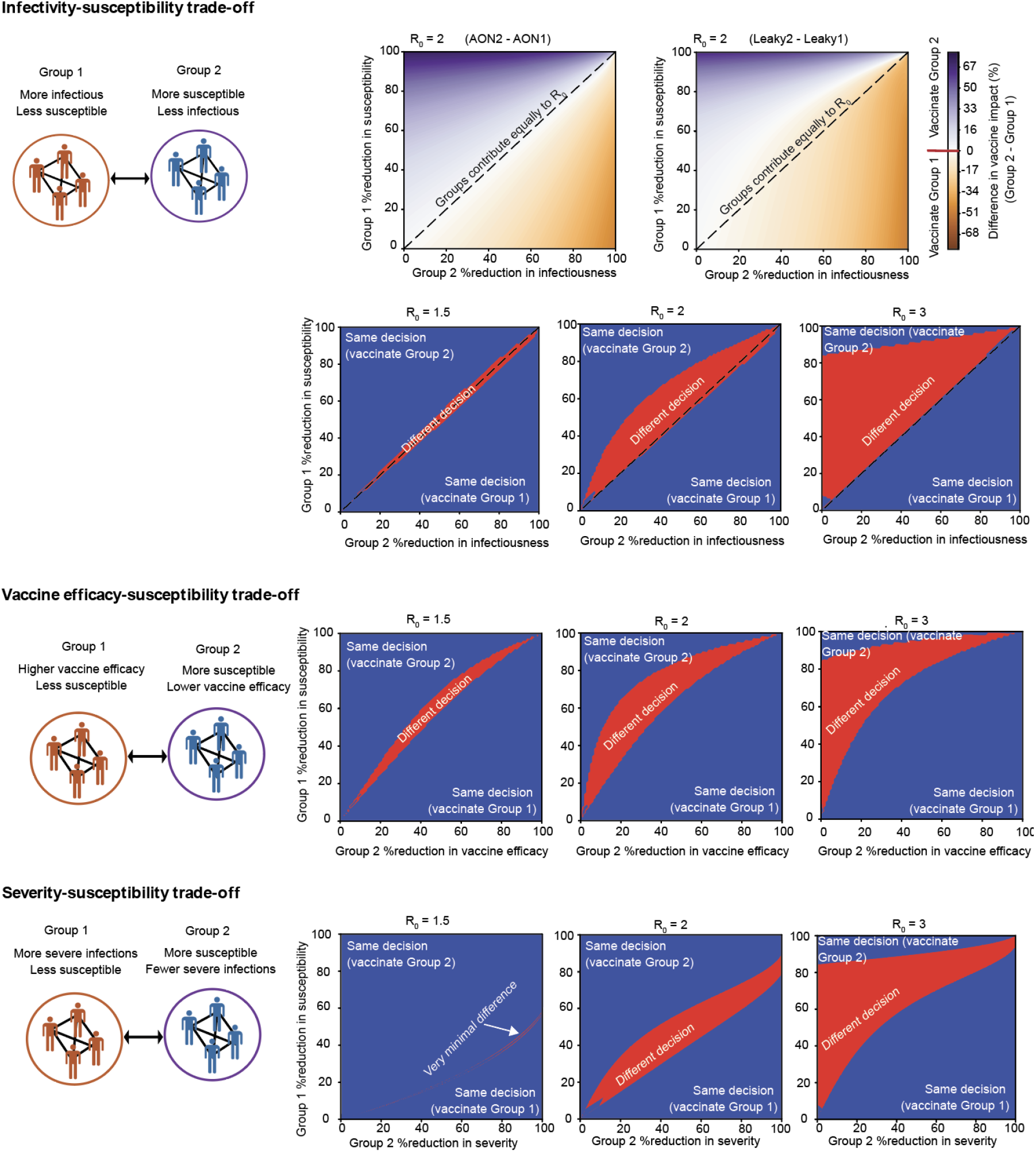
Optimal vaccine allocation strategy depends on mode of vaccine failure. Plots show the difference in vaccine impact between vaccinating Group 2 (more susceptible) vs Group 1 (more infectious, higher vaccine efficacy, or higher risk of severe infection) as a function of the degree of the differences in susceptibilities between the two groups, for a fixed pre-vaccination R_0_, vaccine coverage (50%) and vaccine efficacy (80%). The optimal vaccine allocation decision (“vaccinate Group 1” vs “vaccinate Group 2”) is the one that maximizes the vaccine impact. The heatmap color either shows the magnitude of the difference in predicted vaccine impact between the decisions to allocate vaccine to either only Group 1 or only Group 2 (purple/orange scale), or indicates regions in the parameter space where the optimal vaccination strategy is the same (blue) or different (red) for the two modes of vaccine failure. See Methods for more details.

In our first scenario, one group has higher infectiousness (propensity to transmit if infected) but the other is more susceptible to infection (Figure 6). As expected, we find that the optimal vaccination strategy is parameter dependent, but we also observe that the two models of vaccine failure can lead to conflicting conclusions. There are regions of parameter space where if we assume vaccine-induced protection is all-or-nothing, the optimal strategy is to vaccinate the more susceptible group, but if we assume leaky protection, the less susceptible/more infectious group should be vaccinated. These are not always regimes where the difference between the two vaccine allocation strategies is minimal - sometimes the difference in predicted vaccine impact between the two allocation strategies can be ∼20%. These conflicting model conclusions occur in parameter regimes where the relative differences in infectiousness and susceptibility are similar between the two groups, such that each contributes similarly to overall R_0_ in the absence of vaccination. The region of discrepancy gets amplified with higher overall R_0_ values). These results hold qualitatively for other values of the vaccine efficacy and coverage, but the effect of increasing R_0_ is not as pronounced for vaccines with a higher efficacy (Suppl. Fig. S2).

We can understand this finding as follows. When assuming all-or-nothing protection, the optimal strategy is determined by which of the two groups contributes more to R_0_ in the absence of vaccination (diagonal line separates blue and orange regions in the top line of Figure 6, see SI for details). Since all-or-nothing protection implies that some vaccinated individuals are fully protected, vaccinating the group with the higher contribution to R_0_ is always the best strategy to lower the number of breakthrough infections and thus overall epidemic size. However, this simple rule isn’t necessarily true for leaky vaccines. If the overall force of infection is large enough, it may not be optimal to vaccinate the more susceptible group even when they contribute more to R_0_. These more-susceptible-yet-vaccinated individuals may still have enough remaining susceptibility to experience breakthrough infections due to multiple exposures. In such situations, vaccinating the less susceptible group is more favorable since post-vaccination susceptibility - and thus breakthrough infections - are lower. Said another way, when vaccine protection is leaky, it can sometimes be better to vaccinate a relatively unimportant group if doing so would effectively eliminate their contribution to transmission - thus creating something akin to the perfectly protected group that results from administration of vaccines with all-or-nothing protection.

We considered two other scenarios that would lead to ambiguities in which risk group would be optimal to vaccinate (Figure 6, bottom two rows). In one example, the vaccine is more efficacious against infection for one group, but the other group is more susceptible to infection. In another example, one group has a higher chance of developing severe disease if infected, but the other is more susceptible to infection. In this scenario, vaccinating the more susceptible group will always produce fewer cumulative infections, but the metric for an optimal vaccine becomes the prevention of severe infections. We again find conflicting predictions for the optimal vaccine allocation strategy depending on the way vaccine failure is modeled. In some parameter regimes, especially when baseline R_0_ is high, the leaky model of vaccine protection would suggest it would be optimal to vaccinate the more susceptible group, but the alternative all-or-nothing model of vaccine efficacy would instead conclude that it would be optimal to vaccinate the group more at risk for severe outcomes or for which the vaccine is more efficacious. We also considered scenarios where the groups don’t differ in susceptibility, but face other tradeoffs - for example, one group has higher infectiousness while the other has higher vaccine efficacy. We found there is rarely a discrepancy in predictions of the optimal vaccination strategy between models of leaky vs all-or-nothing vaccines in these cases (Suppl. Fig. S3).

## Discussion

Mathematical models are a critical tool for predicting and evaluating the impact of interventions like vaccination to control infectious diseases. In constructing models, a set of common but often poorly articulated assumptions revolve around how observed population-averaged parameters are distributed across individuals; for example, the distribution of individual-level infectivity, contact patterns, duration of infectiousness, susceptibility, or intervention efficacy. Here we show that when modeling vaccines that reduce infection risk during epidemics, the choice between choosing to model imperfect vaccines as having “leaky” vs “all-or-nothing” efficacy can have a significant impact on model predictions. Assuming “all-or-nothing” protection - in which a vaccine either offers complete or no protection - always results in more optimistic predictions of population-level vaccine impact relative to models where the vaccine is assumed to lead to a uniform per-exposure risk reduction across vaccine recipients (“leaky” protection). This difference is more extreme for outbreaks spreading with higher basic reproduction numbers, for vaccines with intermediate efficacy, and for vaccine campaigns that slow spread without curtailing it entirely, and can lead to differences of up to 30% in calculations of infections averted by vaccination. Using highly simplified models, we highlighted how this effect could have affected predictions made during COVID-19, concluding that it was likely to be largest when estimating residual vaccine impact during the emergence of the Omicron variant at the end of 2021. Our overall conclusion is that the choice of how to model vaccine failure mode is a modeling decision that should be clearly disclosed and ideally included as an axis of uncertainty in model output, just as uncertainty in the value of specific parameters or other aspects of model structure would be.

One of the more common use-cases for mathematical models of vaccines is to compare strategies for allocating limited vaccines among different risk groups - for example, those who contribute more to onward transmission versus those who are at higher risk of severe outcomes if infected; or the very young versus the very old. The qualitative predictions of these scenario comparison analyses, i.e. “prioritize vaccinating Group A over Group B”, are generally assumed to be more robust to specific model assumptions than quantitative predictions about specific cases averted. However, we show here that the choice of how to model vaccine failure mode can sometimes reverse the conclusions about optimal allocation. Furthermore, we note that even if the two models of vaccine failure agree on the optimal vaccination strategy, using the wrong model can result in substantial under or over estimation of the vaccine impact, especially if the optimal strategy involves vaccinating the more susceptible group (Figure S4). For example, in the scenario where groups differ in infectiousness vs susceptibility, we find that even when both models agree that the optimal vaccination strategy is to vaccinate the more susceptible/less infectious group, using an all-or-nothing model of vaccine failure when in fact the vaccine failure mode was leaky can lead to more than 20% overestimation in the vaccine impact (Figure S4C). Therefore, models should specifically evaluate the robustness of their conclusions to their underlying assumptions about vaccine failure mode.

There are several important assumptions to keep in mind with our analysis. We only examined two extreme models for vaccine failure - identical (but imperfect) protection in all vaccinated individuals (leaky), versus an extreme binary distribution of protection (all or nothing). In reality, vaccine effects may be somewhere in between; for example, some individuals may get no protection but those who are protected may have a more complex distribution of protection levels. We haven’t considered that the vaccine failure model could differ between subgroups in the population, for example by age, immune status, pathogen strain, or prior infection history. Our model doesn’t include any other individual-level heterogeneities, such as baseline variation in the degree of susceptibility or infectiousness. We did not examine another dimension of vaccine failure - waning of protection levels over time since vaccination - or models that combine failure via waning with failure in leaky versus all-or-nothing models. The leaky versus all-or-nothing dichotomy may also exist for infection-induced immunity, but in our simple models we have assumed natural immunity is complete in all individuals. Ragonnet *et al*. found that the degree of protection conferred by prior infection modulated the difference between vaccine impact predicted by leaky vs all-or-nothing models for endemic infections, concluding - like us - that the main driver of differences was the number of breakthrough infections post-vaccination, but they did not explore the *distribution* of this infection-induced immunity [15]. Prior theoretical and experimental studies have found that one of the more significant manifestations of the difference between leaky vs all-or-nothing vaccines is in the relationship between exposure dose (i.e., inoculum size) and infection risk [19,42]. Consequently, the importance of differences between these vaccine failure modes could vary between settings and over time depending on factors that modulate this dose, such as use of antimicrobial therapy, use of personal protective equipment like masks, behavior changes, or general health status.

The reason that imperfect vaccines are typically assumed to fail by either “leaky” or “all-or-nothing” modes is because generally it is impossible to measure the distribution of vaccine effects across individuals, especially on the timescale in which mathematical models are needed for decision making. Measuring the distribution of protection levels requires a setting where either the number of exposures or size of exposure doses can be quantified in relation to protection [8,12,18–20,42–46]; the relationship between either exposure metric and the proportion of the population infected can be analyzed mathematically to extract the distribution of susceptibilities. Other than challenge studies - which are expensive in animal models and very rarely ethically suitable in humans - there are few situations in which such a relationship can be obtained. A recent study of COVID-19 risk in vaccinated prisoners used the co-location of infected individuals with their recently-infected contacts (shared cell, same cell-block, or none) as a proxy for exposure intensity, and the observed relationship suggested a leaky mode of vaccine failure [47]. Even if there is an immune marker inferred or strongly suspected to be a correlate of protection, the distribution of this marker does not necessarily need to match the distribution of the degree of protection [9]. For example, a unimodal, continuous distribution of neutralizing antibody levels across a population could still be consistent with all-or-nothing protection if the relationship between antibody level and per-exposure probability of infection is step-like. Zachreson *et al*. used observed relationships between cohort-level neutralizing antibodies and protection against COVID-19 in vaccine trials [48] to propose a consistent model of individual-level protection, showing that when combined with waning antibody levels over time, complex distributions in between the leaky and all-or-nothing extremes can arise [49].

Our results build upon prior work over the past 50 years showing that the distribution of vaccine effects across individuals will impact estimates of vaccine efficacy in clinical or field trials [8,10,11] and model predictions of infections averted by vaccines [12–15,50]. We focus on the context of vaccines administered to the general population during an outbreak (as opposed to routine childhood vaccinations for endemic infections) and systematically evaluate parameter regimes where the choice of failure mode has the largest impact on predictions. We also explicitly focus on when predictions of optimal allocation decisions can be sensitive to the mode of vaccine failure, which some prior studies have done but only in a disease/scenario specific context [25,26,28]. Our observation that all-or-nothing vaccines always provide equal or greater population-level protection is in agreement with other models [12–15], and a prior study of childhood vaccination for endemic diseases found a similar relationship with vaccine efficacy, transmissibility, and likelihood of breakthrough infections when examining how the post-vaccination endemic equilibrium varied with failure mode [15].

Theoretically, since vaccine failure mode impacts disease dynamics, it should be possible to infer failure mode from comparing model predictions to data if there is enough certainty about other model parameters. To understand the persistence of pertussis (whooping cough) transmission despite decades of using vaccines with high efficacy, Domenech de Celles used detailed age-stratified time-series data on pertussis cases before and after vaccination to show that models assuming some degree of “all-or-nothing” failure combined with waning over time were best supported by the data, as opposed to a model of leaky vaccine failure [50]. Prior modeling work by this group showed that vaccine failure mode affects the average age of infection in models of childhood vaccination for endemic diseases, making this identification possible [14]. More generally, retrospective modeling of vaccination campaigns could assist in identification of the failure mode, as long as other important disease mechanisms are reasonably well understood, like any major risk-group specific difference in susceptibility, infectiousness, or vaccine efficacy, as well as the role of waning vaccine-induced protection or antigenic drift.

For prospective vaccine impact modeling studies, our results imply that the vaccine failure model should be included as an axis of uncertainty. In addition, these findings highlight the benefit of clinical trials designed with the longitudinal follow-up required to differentiate vaccine failure modes [11], and suggest that ethical, human challenge studies that could allow identification of the failure model should be considered [51,52].

## Data Availability

This is a mathematical modeling study. All code used to generate results is open-source and available on Github.

https://github.com/diane-lee-01/VaccineEfficacy

## Supplement

### Supplementary Methods

#### Derivation for number of breakthrough infections for the two models of vaccine failure for a simple infection model

To provide intuition behind why the number of breakthrough infections for a leaky vaccine are always equal to or greater than those for an all-or-nothing (AON) vaccine (with the same efficacy and coverage), we consider a simplified infection model with a constant force of infection and compare the number of breakthrough infections under the two models of vaccine failure. Suppose every unvaccinated individual has probability *p* of getting infected per exposure. For a leaky vaccine with efficacy *v*_*e*_, the probability that a vaccinated individual is infected given *n* (≥ 1) exposures is given by,

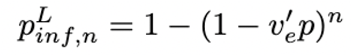

where *v*′ _*e*_ = (1 − *v*_*e*_) and 0 ≤ *v*′ _*e*_ < 1. If *N*_*v*_ individuals are vaccinated then the number of breakthrough infections after *n* exposures is given by,

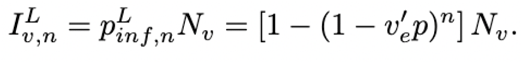

For an AON vaccine, the probability that a vaccinated individual is infected given n exposures is either 0 (for those fully protected) or

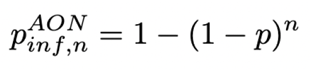

for those still susceptible after vaccination. As a result, if *N*_*v*_ individuals are vaccinated, the number of breakthrough infections after *n* exposures is given by,

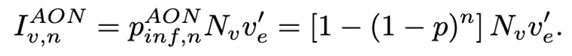

First consider the two limits of exposure: *n* = 1 and *n* → ∞. When *n* = 1,

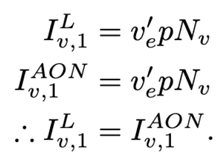

And when *n* → ∞,

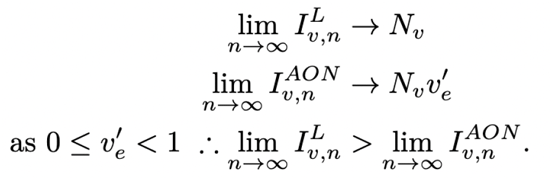

Finally, using the series expansion for (1 − *x*) around zero, for a general number of exposures *n*, the number of breakthrough infections is given by,

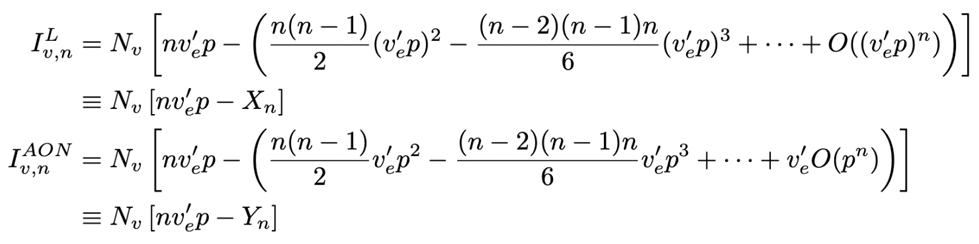

Comparing the two expressions term-by-term we see that after the first term which is shared by both, each of the following terms in *I*^*L*^ _*v,n*_ (i.e., in *X*_*n*_) are smaller or at most equal to the corresponding terms in *Y*_*n*_ as they have higher powers of *v*′ _*e*_ and 0 ≤ *v*′ _*e*_ < 1. Therefore, *X*_*n*_ ≤ *Y*_*n*_ for all *n*, which implies that *I*^*L*^ _*v,n*_ ≥ *I*^*AON*^ *v,n*, that is, the number of breakthrough infections (given a fixed number of exposures) for a leaky vaccine are always greater than or equal to an all-or-nothing one.

### Parameters for applications to COVID-19 vaccine impact

We obtained data on vaccine coverage (defined here as % of individuals who received at least one dose of vaccine), case incidence (daily reported cases per million individuals in the population, 7 day rolling average, right aligned), and real-time effective reproduction number from Our World In Data [3,4]. We identified the variants responsible for the phase of the epidemic of interest from CoVariants.org [32].

#### Scenario: Rapid control of epidemic wave via concurrent vaccination - Alpha wave in fall 2020/winter 2021 in Israel

We defined this wave in Israel as starting Nov 15 2020, peaking around Jan 15 2021, and concluding around March 15 2021. Vaccination began for the first time in mid December, with around 20% vaccinated per month until reaching ∼ 60% vaccinated by the conclusion of the wave. Estimating the proportion of the population with immunity from prior infection at the time of this wave was more complicated than in the US setting, as recurring national serosurveys were not conducted in Portugal. However, Reicher et al estimated seroprevalence in Sept 2020 of 4.5%, and a seroprevalence-to-detected-case ratio ∼4.5 which had decreased over time [53]. Between this time (which was within an smaller pre-Alpha wave) and the start of the Alpha wave in mid November, cumulative incidence of reported cases increased from ∼1.7% about 3.6%, a 1.9% increase, suggesting an increase in seroprevalence of around 8.5% (with ratio of 4.5 - assuming no further improvement in case detection, so likely an overestimate). We thus approximated immunity from prior infection at the start of the Alpha wave as 10%. During the period of exponential growth throughout Dec 2020, R_eff_ varied from 1.25 - 1.4, which with 10% prior immunity (and no vaccination), corresponds to around R_0_ between 1.4-1.55, which we approximated as R_0_ = 1.5.

#### Scenario: Pre-outbreak vaccination with high efficacy - Delta wave in summer 2021, United States

We defined this wave as starting July 1, 2021, peaking in early September, and continuing through the end of October. On July 1 2021, 48% of the population was fully vaccinated and 54% had at least one dose, so we approximated vaccine coverage (f_V_) as 50%. We assumed no extra vaccines occurred during the outbreak - although in reality vaccine coverage increased to 58% fully vaccinated and 65% with at least one dose by Nov 1 2021. Vaccine efficacy (VE) was approximated as 80%, based on estimates of mRNA vaccine efficacy against infection for the Delta variant [29], which dominated during this wave. We abstained data on cumulative prior infections (f_R_) from a seroprevalence survey measuring presence of the SARS-CoV-2 nucleocapsid protein (to differentiate from immunity due to vaccination), collected from a national sample of blood donors [30,31]. During the June 2021 round of sampling, 20.6% of individuals were estimated to be seropositive nationally, so we approximated immunity from prior infection at 20%. During the period of exponential growth centered around Aug 1, 2021, R_eff_ peaked at 1.5. To estimate R_0_ in the absence of immunity from prior infection and vaccination, we used the formula R_eff_ = R_0_*(1-f_R_-f_V_*VE). We assume some overlap between prior infection and vaccination based on the assumption that vaccination status is independent of prior infection status. Thus, if vaccine coverage is 50%, we expect that 10% of that occurs in those with immunity from prior infection (which is assumed to be perfect protection), so only 40% is in previously uninfected individuals. Thus, R_eff_ ∼ R_0_*(1-0.2-0.4*0.8) = R_0_*0.48, so R_0_ = 3.1. We thus approximated R_0_ = 3 during this wave.

#### Scenario: Pre-outbreak vaccination with intermediate efficacy - Omicron wave in fall 2021/winter 2022 in countries with widespread mRNA vaccine coverage (e.g. Portugal)

We defined this Omicron-variant-driven wave in Portugal as starting on Dec 1 2021, peaking late January, and continuing until early March. On Dec 1, 81% of the population was fully vaccinated and 89% had at least one dose; these increased to 84% and 93% by March. We assumed 90% coverage (f_V_) for the duration of the wave. Vaccine efficacy (VE) was roughly approximated as 50% for this period, based on estimates of mRNA vaccine efficacy against infection for the Omicron variant (in reality, studies give a wide range of estimates, and find strong dependence on time since last dose and whether a booster dose was received [33–37]). Due to the lack of protection against infection with Omicron conferred by prior infection with pre-Omicron variants, we assumed effectively 0% of the population had immunity from prior infection. During the period of exponential growth between Dec 1 2021 and Jan 15 2022, the effective reproduction number varied between 1.25 to 1.55, so using R_eff_ = R_0_*(1-f_R_-f_V_*VE) ∼ R_0_*(1-0-0.9*0.5) we get a range for R_0_ of 2.3 - 2.8. We chose R_0_ = 2.3 which gave approximately 10% of the population infected each month during the first few months. Note that a similar phenomenon happened in Singapore, but later - from around mid January to early May 2022.

### Model for exploring vaccine allocation decisions

We extend the SEIRV type model introduced in the main text to include two equal sized groups of individuals. The two groups can differ in their susceptibilities α_*Si*_, infectivities α_*Ii*_, they can have different vaccine efficacies ε_*Ai*_ (all-or-nothing) and ε_*Li*_ (leaky), and different fraction of individuals vaccinated *f*_*vi*_. The vaccine is modeled as instantaneous and is administered at time *t*_*v*_ into the epidemic. This system is described by the following set of ODEs,

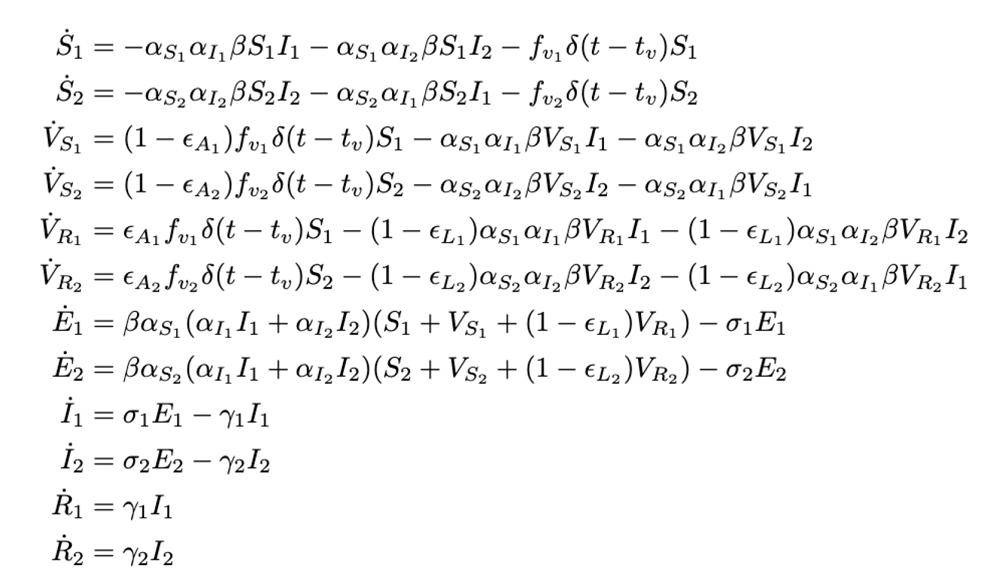

where the state variables *Si, V*_*Si*_, *V*_*Ri*_, *E*_*i*_, *I*_*i*_ and *R*_*i*_ are proportions of the population. For simplicity we assume σ_1_ = σ_2_, γ_1_ = γ_2_ and *t*_*v*_ = 0. R_0_ can be calculated using the next-generation matrix method [54] and is given by,

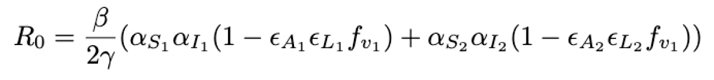

### Parameters used for exploring vaccine allocation decisions

For all scenarios we fix the average duration of latency (σ) and the average duration of infectiousness (γ) to 4 days and vary the other parameters while keeping pre-vaccination R_0_ ∈ {2, 3} fixed.

#### Infectivity susceptibility trade-off (Group 1 is more infectious and Group 2 is more susceptible)

Since Group 1 is more infectious, we assume that the infectivity of Group 2 is reduced compared to Group 1 by a factor *m*, where *m* is varied between 0 and 1,

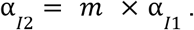

Similarly, as Group 2 is more susceptible, we assume that the susceptibility of Group 1 is reduced compared to Group 2 by a factor *n*, where *n* is varied between 0 and 1,

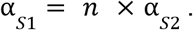

We assume α_*I*1_ = α_*S*2_ = 1. With this parameterization, the pre-vaccination R_0_ for this scenario is given by,

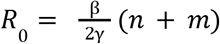

We vary β to keep R_0_ fixed as we vary *m* and *n*.

The following values are used for vaccine efficacies and coverage-levels for the three sub-scenarios (*i* = 1, 2):

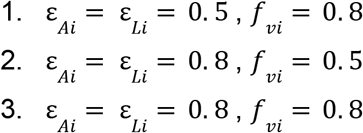

#### Vaccine efficacy and susceptibility trade-off (Group 1 has higher vaccine efficacy and Group 2 is more susceptible)

Since Group 1 has higher vaccine efficacy, we assume that the vaccine efficacy for Group 2 is reduced compared to Group 1 by a factor *n*, where *n* is varied between 0 and 1,

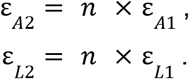

As Group 2 is more susceptible, we assume that the susceptibility of Group 1 is reduced compared to Group 2 by a factor *m*, where *m* is varied between 0 and 1,

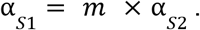

We assume α_*S*2_ = α_*I*1_ = α_*I*2_ = 1. With this parameterization, the pre-vaccination R_0_ for this scenario is given by,

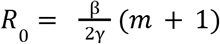

We vary β to keep R_0_ fixed as we vary *m*.

The following values are used for vaccine efficacies for Group 1 (Group 2’s are varied as above) and coverage-levels (for both groups) for the three sub-scenarios (*i* = 1, 2):

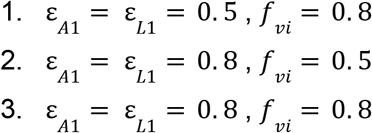

#### Disease severity and susceptibility trade-off (Group 1 more severe infections and Group 2 is more susceptible)

Since our model doesn’t explicitly consider different stages of infection severity, we assume that at 1% of infections in Group 1 lead to severe disease. This is then reduced for Group 2 by a factor *m*, where *m* is varied between 0 and 1.

As for the first scenario, as Group 2 is more susceptible, we assume that the susceptibility of Group 1 is reduced compared to Group 2 by a factor *n*, where *n* is varied between 0 and 1,

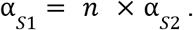

We assume α_*I*1_ = α_*I*2_ = α_*S*2_ = 1. With this parameterization, the pre-vaccination R_0_ for this scenario is given by,

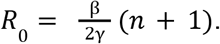

We vary β to keep R_0_ fixed as we vary *n*.

The following values are used for vaccine efficacies and coverage-levels for the three sub-scenarios (*i* = 1, 2):

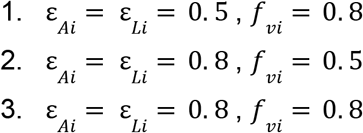

## Supplementary Figures

**Figure S1:**
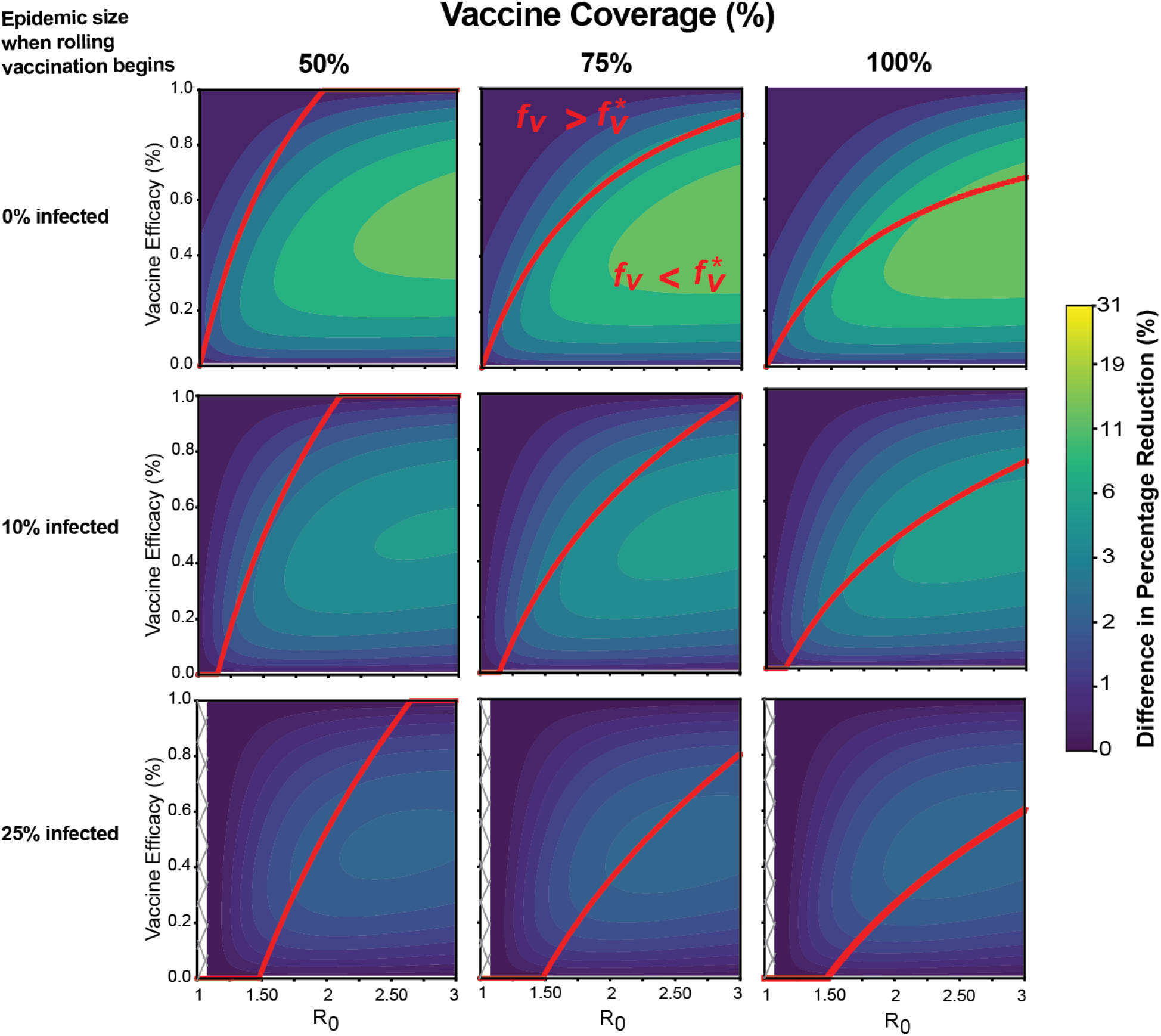
Difference in impact between all-or-nothing and leaky vaccines for rolling vaccine administration. Difference in vaccine impact for different levels of vaccine coverage and timing of vaccination over a range of vaccine efficacies and R_0_ values for fixed rate of daily vaccination rate (5% of the susceptible unvaccinated population). The colorbar indicates the difference in the percentage reduction of the final epidemic size compared to that of without vaccination between leaky and all-or-nothing vaccines. Red line separates regions in the parameter space where the fraction vaccinated are greater (above) or less (below) than the herd immunity threshold. Uncolored hashed regions correspond to parameter combinations where disease R_0_ was too small for the epidemic to reach the target size for vaccine administration (so vaccine was never administered). See Methods for more details on how rolling vaccination was implemented.

**Figure S2:**
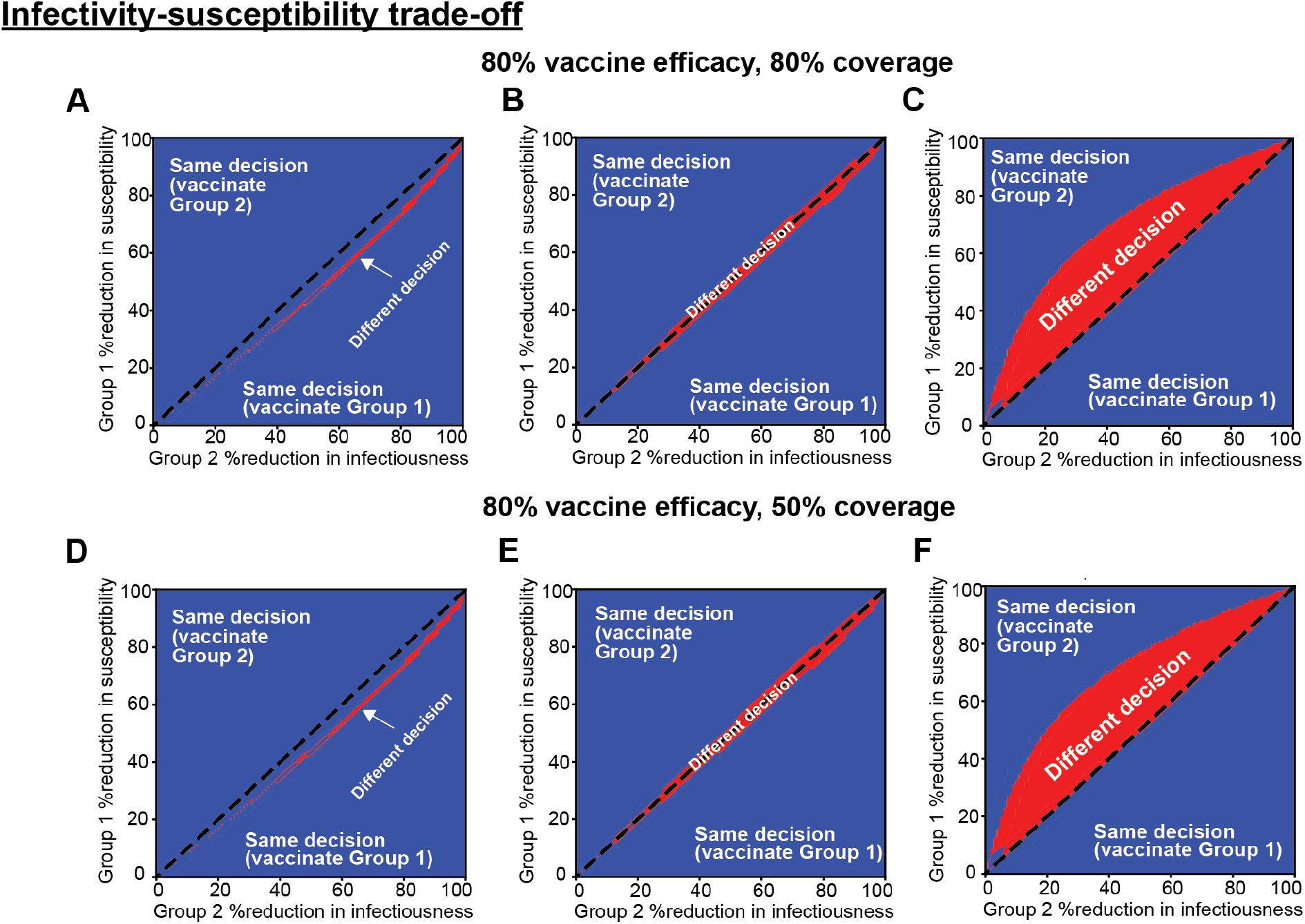
Effect of the two modes of vaccine failure on optimal vaccine allocation decisions. Results for scenario where Group 1 is more infectious than Group 2 but Group 2 is more susceptible than Group 1 for an all-or-nothing or leaky vaccine with 80% efficacy and A)-C) 80% or D)-F) 50% coverage. Plots show regions in the parameter space on varying infectiousness and susceptibility where the optimal vaccination strategy is the same (blue) or different (red) for the two modes of vaccine failure for three different values of pre-vaccination R_0_.

**Figure S3:**
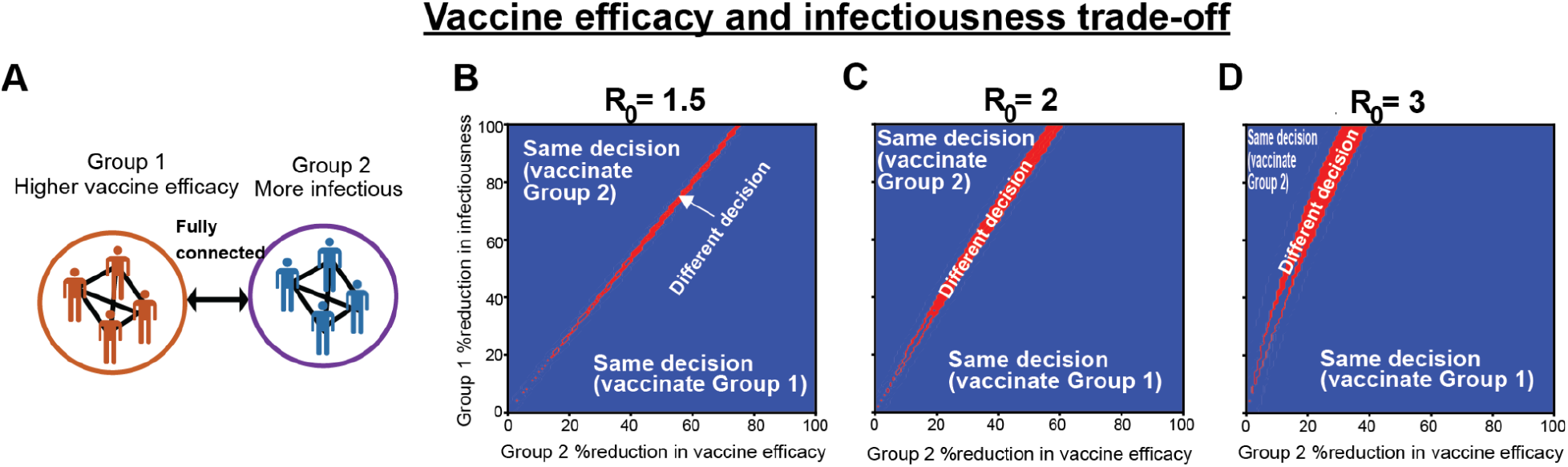
Effect of the two modes of vaccine failure on optimal vaccine allocation decisions. Results for scenario where vaccine efficacy is higher for Group 1 than Group 2 but Group 2 is more infectious than Group 1. B)-D) Regions in the parameter space on varying infectiousness and vaccine efficacy where the optimal vaccination strategy is the same (blue) or different (red) for the two modes of vaccine failure for fixed vaccine coverage (50%) for three different values of pre-vaccination R_0_.

**Figure S4:**
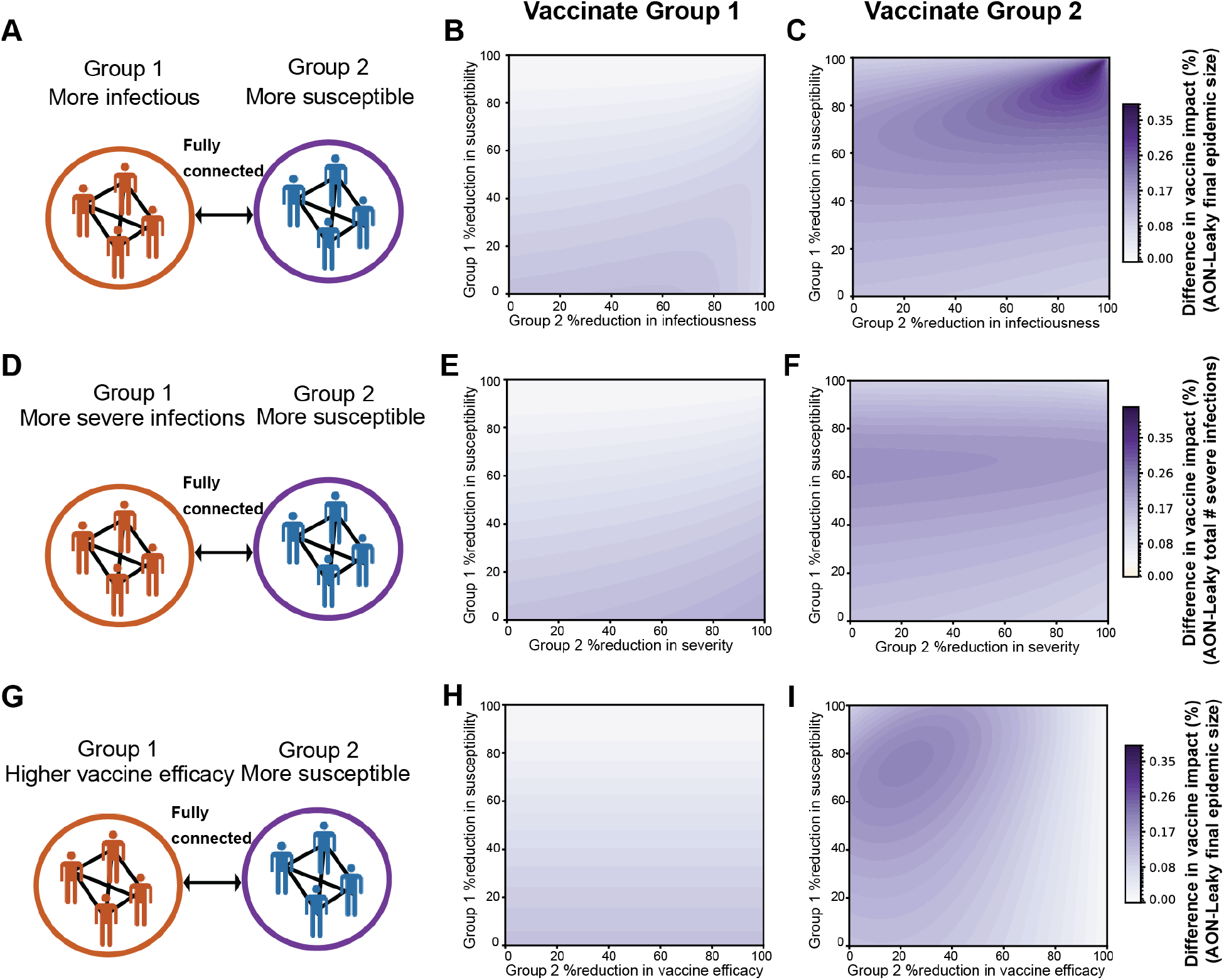
Difference in vaccine impact on vaccinating either of the groups with AON or leaky vaccines. A)-C) Results for scenario with infectiousness and susceptibility trade-off. Difference in vaccine impact on vaccinating B) Group 1 or C) Group 2 with all-or-nothing or leaky vaccines. The colorbar indicates the difference in the percentage reduction of the final epidemic size compared to that of without vaccination between all-or-nothing and leaky vaccines. (Middle row) Same as above for a scenario with disease severity and susceptibility trade-off but the colorbar indicates the difference in the percentage reduction of the final number of severe infections compared to that of without vaccination between all-or-nothing and leaky vaccines. (Bottom row) Results for scenario with vaccine efficacy and susceptibility trade-off, with the same metric of vaccine impact as in the top row. Results are for baseline *R*_0_ = 2, 50% baseline vaccine efficacy and 80% coverage.

